# Exploring Heteroplasmic Variants in mtDNA: Insights from Single-Cell Transcriptomics

**DOI:** 10.64898/2026.05.06.26352529

**Authors:** Marco Barresi, Ivano Di Meo, Alessia Nasca, Eleonora Lamantea, Andrea Legati, Daniele Ghezzi

**Author notes:** **Correspondence:** Daniele Ghezzi.

## Abstract

Mitochondrial DNA (mtDNA) heteroplasmy, which is the coexistence of wild-type and mutant mtDNA variants within the same cell, plays a critical role in modulating cellular phenotype as well as disease severity and penetrance. Bulk RNA sequencing is not able to detect cell-to-cell variability in heteroplasmy, limiting our understanding of mitochondrial pathological mechanisms. In this study, we leverage single-cell RNA sequencing (scRNA-seq) combined with a robust bioinformatics pipeline to characterize mtDNA heteroplasmy. We employed four fibroblast lines from patients harboring heteroplasmic mtDNA pathogenic variants in genes encoding respiratory complex I subunits. While RNA heteroplasmy corresponds to DNA-based measurements at the bulk-level, single-cell analysis uncovers a diverged distribution: most cells have near-homoplasmic (wild-type or mutant) mtDNA, with few cells showing intermediate levels. Furthermore, we find that high mutation levels correlate with transcriptional profile changes, though these responses are highly sample-specific, suggesting that nuclear background and cellular context critically influence mitochondrial dysfunction and compensatory mechanisms. Our findings highlight the power of single-cell technologies to better understand the complex link between mtDNA genetic diversity and mitochondrial phenotypic variability, and to study crucial aspects in mitochondrial biology and pathology, such as clonal dynamics, at single-cell resolution.

## 1 Introduction

The study of mitochondrial heteroplasmy, defined by the coexistence of wild-type and acquired mutant variants of mitochondrial DNA (mtDNA), is crucial for understanding significant biological processes and pathologies associated with mitochondrial damage or mtDNA dysfunction. Each cell contains hundreds/thousands of mtDNA copies, and the relative frequencies of heteroplasmic variants can vary considerably among individual cells, even within the same tissue, thereby profoundly influencing cellular phenotype, suggesting dynamic processes of acquisition and selection of mitochondrial variants (Nissanka and Moraes, 2020; Parakatselaki and Ladoukakis, 2021; Stewart and Chinnery, 2021). It is well established that the extent of heteroplasmy modulates mitochondrial functional impairment, with effects determined by both the mutational threshold and the specific nature of the mutation. Elucidating the distribution and functional consequences of these mtDNA variants is therefore critical for understanding the pathogenic mechanisms underlying a wide range of mitochondrial, neurodegenerative, and systemic diseases, as well as for advancing knowledge of mitochondrial biology under physiological conditions.

Bulk RNA sequencing has long been established as the primary method for analysing mtDNA variants and assessing their effect on transcriptional profile, which provides an average estimate of allelic composition. However, this approach doesn’t account for the considerable variability in heteroplasmy among individual cells: the same average percentage of variant measured in bulk can arise from very different distributions at the cellular level (e.g., many cells with intermediate heteroplasmy vs. a mix of nearly homoplasmic wild-type or mutant cells) (Cloonan et al., 2008; Payne et al., 2013). This level of variability is crucial for interpreting the penetrance and phenotypic variability of mitochondrial diseases, as well as for understanding processes such as clonal selection and mitochondrial genetic drift.

Single-cell technologies (scRNA-seq, scATAC-seq, and multimodal approaches) have transformed investigations into this genomic and transcriptional heterogeneity, enabling precise characterization of mtDNA variants at single-cell resolution and rigorous assessment of their effects on cellular function (Marshall and Jones, 2021; Nitsch et al., 2024). The biological relevance of these technologies has been repeatedly demonstrated in studies for lineage tracing and investigations of mtDNA segregation dynamics, both in vitro and in vivo, across diverse cell types and disease contexts (Ludwig et al., 2019). Various single-cell datasets have been used to reconstruct clonal relationships by exploiting mtDNA variants as endogenous genetic barcodes and map the distribution of heteroplasmic variants within tissues, revealing associations between specific mtDNA genotypes and defined cellular subpopulations. These studies also highlight how mtDNA mutations can modulate mitochondrial network dynamics and influence cellular gene-expression programs.

Accurate estimation of heteroplasmy at single-cell resolution remains challenging (Lareau et al., 2023). Measurements can be confounded by sequencing errors, PCR and library-preparation amplification biases, uneven coverage, and misassignment of reads deriving from nuclear mitochondrial DNA segments (NUMTs). Many of these limitations have been partially mitigated by recent advances - both experimental (e.g., targeted mitochondrial enrichment of scRNA libraries and full-length single-cell protocols) and computational (dedicated mtDNA variants pipelines) - which integrate targeted enrichment and robust statistical and bioinformatics frameworks in order to obtain heteroplasmy estimates at cell-level suitable for downstream functional inference (Miller et al., 2022; Nakagawa et al., 2025).

In this context, our study aims to expand current understanding of mtDNA heteroplasmy by combining scRNA-seq with an integrated bioinformatics workflow to characterize the cellular distribution of known heteroplasmic mtDNA variants at single-cell resolution in patient-derived fibroblast lines and evaluate the impact of variant heteroplasmic percentage on global transcriptional programs.

## 2 Materials and Methods

### 2.1 scRNA-seq library preparation and sequencing

Primary skin fibroblasts from four patients carrying heteroplasmic variants primarily located in the NADH dehydrogenase (ND) genes – m.11777C>A (MT-ND4), m.13513G>A (MT-ND5), m.13514A>G (MT-ND5), and m.13063G>A (MT-ND5) - were used in this study. Cells were grown in complete medium composed of Dulbecco’s Modified Eagle Medium high glucose (4.5 g/L), supplemented with 2 mM L-Gln, 100 U/mL penicillin, 10 µg/mL streptomycin, and 10% fetal bovine serum (FBS). Each cell lines were collected and re-suspended in DPBS, and the cell suspensions (> 90% living cells examined by count) were loaded on a Chromium Single Cell Controller (10x Genomics). Single-cell RNA sequencing was performed using the 10x Genomics Chromium Single Cell 3’ kit v3.1 (10x Genomics, Pleasanton, CA, USA). Cells were encapsulated into Gel Bead-in-Emulsions (GEMs) together with barcoded oligonucleotides, enabling reverse transcription, cDNA synthesis, and subsequent library preparation according to the manufacturer’s protocol (Danielski, 2023). The scRNA-seq libraries were sequenced on Illumina NovaSeq 6000 platform with paired-end, dual-indexing reads.

### 2.2 scRNA-seq data processing

Raw sequencing data for all samples were processed by a data analysis workflow implemented utilizing integrated bioinformatics pipelines within Linux-based and R environments built on cluster HPC. Cellranger (v7.1.0) is used with the GRCh38 transcriptome reference to obtain per-cell alignment BAMs files and filtered 10× count matrices. Across the analyzed samples, we obtained approximately 3,000 to 6,500 cells per sample, with mean reads per cell ranging from 161,000 to 230,000. Mitochondrial reads pileups, per-cell allele counts, and per-cell heteroplasmy estimates (allele frequency) were obtained with MitoTrace (v. 1.0.0) (Wang et al., 2023), applied to the Cellranger BAMs files along with reference genomic sequence in GRCh38.fasta format. To limit noise from low-coverage barcodes, we excluded barcodes with fewer than 100 mitochondrial reads (min_read = 100) and considered a site covered in a cell only if local coverage was ≥ 2 reads. Count matrices were converted to Seurat objects (Seurat v. 5.3.0) (Butler et al., 2018). This step removes barcodes with fewer than 100 detected features (thereby filtering potential empty/low-quality droplets) and excludes genes detected in fewer than three cells. Allele frequencies at mtDNA positions of interest were integrated into Seurat objects metadata, and to maximize cell retention for heteroplasmy profiling, we did not apply other filters on total genes detected or mitochondrial read fraction; nevertheless, percent_mito and nFeature_RNA were computed for each cell and regressed out during scaling to mitigate technical confounding. Seurat preprocessing included normalization, identification of 2,000 highly variable features, scaling while regressing nFeature_RNA and percent_mito, and PCA (npcs = 20). The number of PCs retained for downstream embeddings and clustering was determined via elbow plots and cumulative variance explained; for the example dataset, 16 PCs were used. Nonlinear embeddings (UMAP, t-SNE) and clustering (resolution = 0.3) were computed on the selected PCs. Based on the observed cell-level distributions of heteroplasmy, we defined two extreme categories for functional comparisons: low heteroplasmy (0–10%) and high heteroplasmy (90–100%). These categories were stored as a metadata field (HET_category) and used as the condition variable in differential expression analyses, both for per-sample and pooled plate-level comparison.

Single-cell differential expression analysis was performed with MAST statistical hurdle models (v. 1.33.0) (Finak et al., 2015). Seurat objects were converted to SingleCellAssay objects, and counts were log-transformed as log2(counts + 1). Genes expressed in fewer than 20% of cells were excluded from MAST to reduce low-signal noise. For per-sample comparisons, we simply fitted the zlm model with only the heteroplasmy categories condition and tested the contrast high vs low. For the pooled plate-level comparison, we fit a model that included, in addition to the heteroplasmy condition, control covariates for the sample and for the detection of cell rate.

Relevant terms were extracted from the MAST results: hurdle p-values (r(>Chisq)), coefficients (coef), and component estimates for the discrete (β_D) and continuous (β_C) parts of the model. Fold-changes were computed as exp(coef) and expressed also as log2 when reported. An operational selection threshold of Pr(>Chisq) < 0.05 was used to prioritize genes for downstream annotation; false discovery rates (Benjamini–Hochberg) were computed and reported in supplementary tables. To characterize the mechanism underlying expression changes, genes were classified according to the sign and concordance of coef, β_D, and β_C into categories such as Concordant_up, Concordant_down, Discrete_only_driver, Continuous_only_driver, and Discordant_other, thereby distinguishing changes driven primarily by alterations in the fraction of expressing cells from those driven by changes in per-cell expression intensity.

Differentially expressed genes were mapped to Entrez identifiers (org.Hs.eg.db) and subjected to pathway and gene ontology enrichment analyses (clusterProfiler: GO BP/CC/MF, KEGG, Reactome). Mitochondrial gene enrichment was specifically using the Human MitoCarta 3.0 gene list and internal panels; the fraction of DE genes corresponding to MitoCarta genes was reported, and overlaps across samples were visualized (UpSet and Heat-tile plots).

## 3 Results

### 3.1 Mitochondrial transcript coverage in scRNA-sequencing

We focused our analysis on the mitochondrial transcriptome by extracting all reads that mapped to the mtDNA. We evaluated the coverage of the 13 transcripts encoded by the mtDNA and compared it to that obtained by bulk RNA-seq. The coverage was not evenly distributed and is strongly biased toward the 3′ end of transcripts. In contrast, bulk RNA-seq yields deep, more uniform coverage across all mtDNA genes (Figure 1).

**Figure 1.**
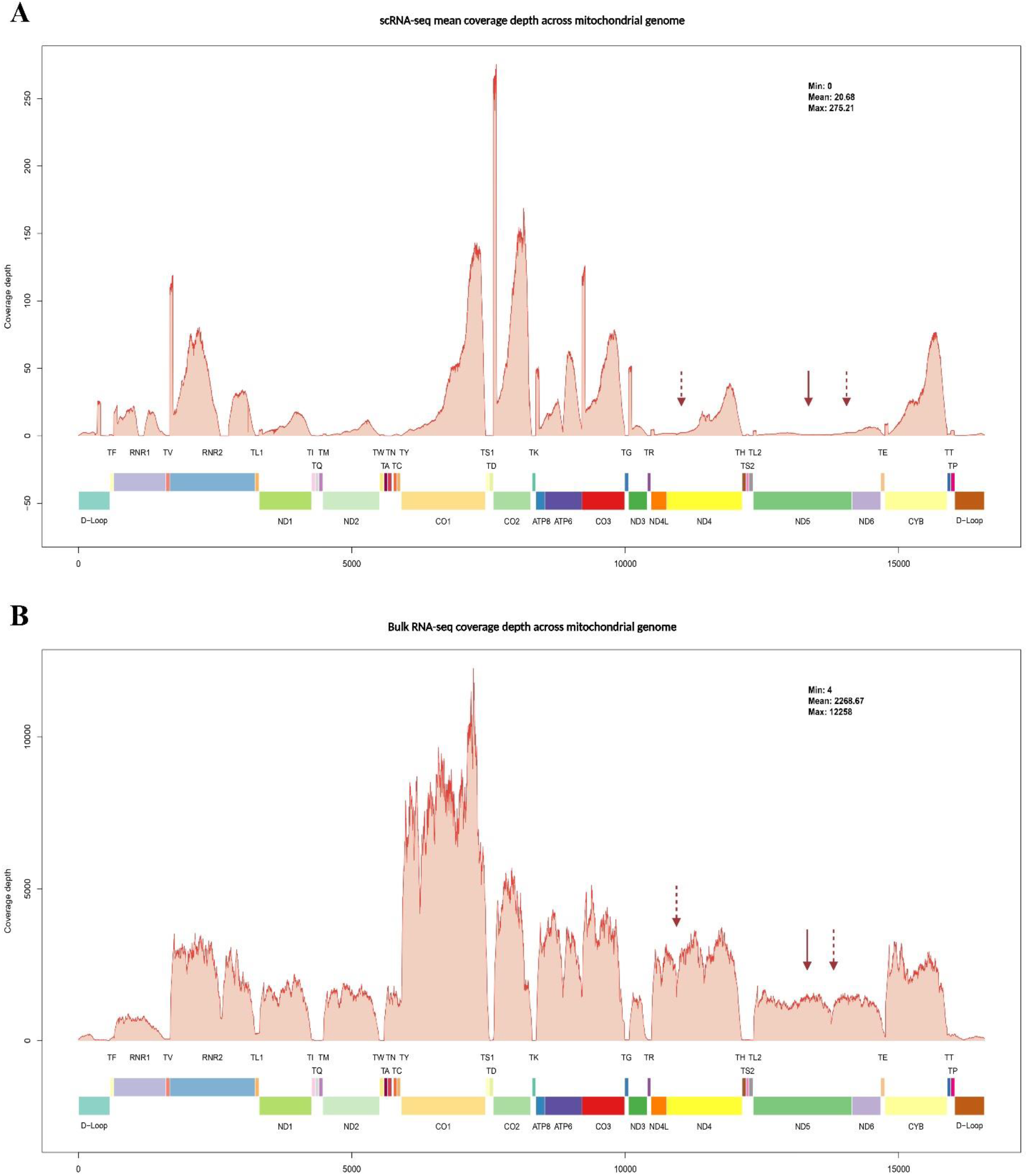
Mitochondrial coverage profiles using scRNA-seq and bulk RNA-seq and pile-up comparison in sample S1 (m.13063G>A, MT-ND5) (A) Mean per-cell coverage across the mitochondrial genome obtained by scRNA-seq (10× Genomics poly-A capture). The solid arrow marks the ND5 variant site in S1; dashed arrows indicate the positions of other mtDNA variants analyzed in this study. (B) Bulk RNA-seq coverage profile with a substantially higher total read count-demonstrating deep, uniform coverage of ND5 comparable to other Complex I subunits.

### 3.2 Heteroplasmy evaluation from scRNA-seq data

Since the analyzed samples harboured heteroplasmic variants in mtDNA, we assessed the mutation load in RNA-seq data. Three samples (S1-S3) had variants in MT-ND5 and one (S4) in MT-ND4. Notably, both the MT-ND5 gene (~1.8 kb) and MT-ND4 (~1.4 kb) exhibit poor, uneven coverage in scRNA-seq data: as reported above, this can be since single-end reads generated from the 3′ terminus rarely reach their central and 5′ regions (Figure 1). Furthermore, MT-ND5 showed the lowest mean depth of coverage amongst mitochondrial transcripts, possibly because it is one of the longest mitochondrial genes. Nonetheless, the coverage, even in these poorly represented transcripts, was enough for an estimation of the RNA heteroplasmy at a single-cell level.

We calculated the mean RNA heteroplasmy from pooled reads across all cells (pseudo-bulk) and compared the result with RNA mutation load detected by bulk RNA-seq or with DNA heteroplasmy. For all the variants, the mean scRNA heteroplasmy was mostly comparable to that identified in bulk RNA and DNA (Table 1).

**Table 1.** Comparison of heteroplasmy levels in genomic DNA and single-cell RNA (scRNA) for four mitochondrial DNA variants.

| Sample | Gene | mtDNA Variant | DNA heteroplasmy | Bulk RNA heteroplasmy | scRNA Heteroplasy* | ScRNA mean coverage |
| --- | --- | --- | --- | --- | --- | --- |
| S1 | *MT-ND5* | m.13063G>A | 38% | 36% | 38% | 5.61 |
| S2 | *MT-ND5* | m.13514A>G | 60% | 64% | 58% | 3.11 |
| S3 | *MT-ND5* | m.13513G>A | 63% | 65% | 69% | 3.11 |
| S4 | *MT-ND4* | m.11777C>A | 68% | 68% | 72% | 34.90 |

\*Heteroplasmy percentages for scRNA were calculated from pooled reads across all cells with at least 2x depth of coverage in the specific heteroplasmic position.

### 3.3 Differences in RNA heteroplasmy among single cells from the same subject

Despite the tested samples had a mean RNA heteroplasmy between 38% and 69%, analysis of per-cell heteroplasmy distribution revealed that S1-S3 samples showed a bimodal distribution with most cells near homoplasmic wild-type (heteroplasmy 0-10%) or homoplasmic mutant status (heteroplasmy 90-100%) and few cells displaying intermediate percentages of mutation load (Figure 2), while S4 showed a more “Gaussian” distribution plus a peak for near homoplasmic mutant status. To assess whether the distinctive distribution observed in S4 reflected altered transcript abundance, we directly compared per-cell abundance of MT-ND4 and MT-ND5 between high-mutant heteroplasmy (90–100%) and low-mutant heteroplasmy (0–10%) populations across all four samples. Neither raw read counts nor log–normalized expression values for MT-ND4 or MT-ND5 differed significantly between the two groups in any patient line (all FDR-adjusted p-values > 0.4; Tables S1-2), including S4 (Figure S1).

**Figure 2.**
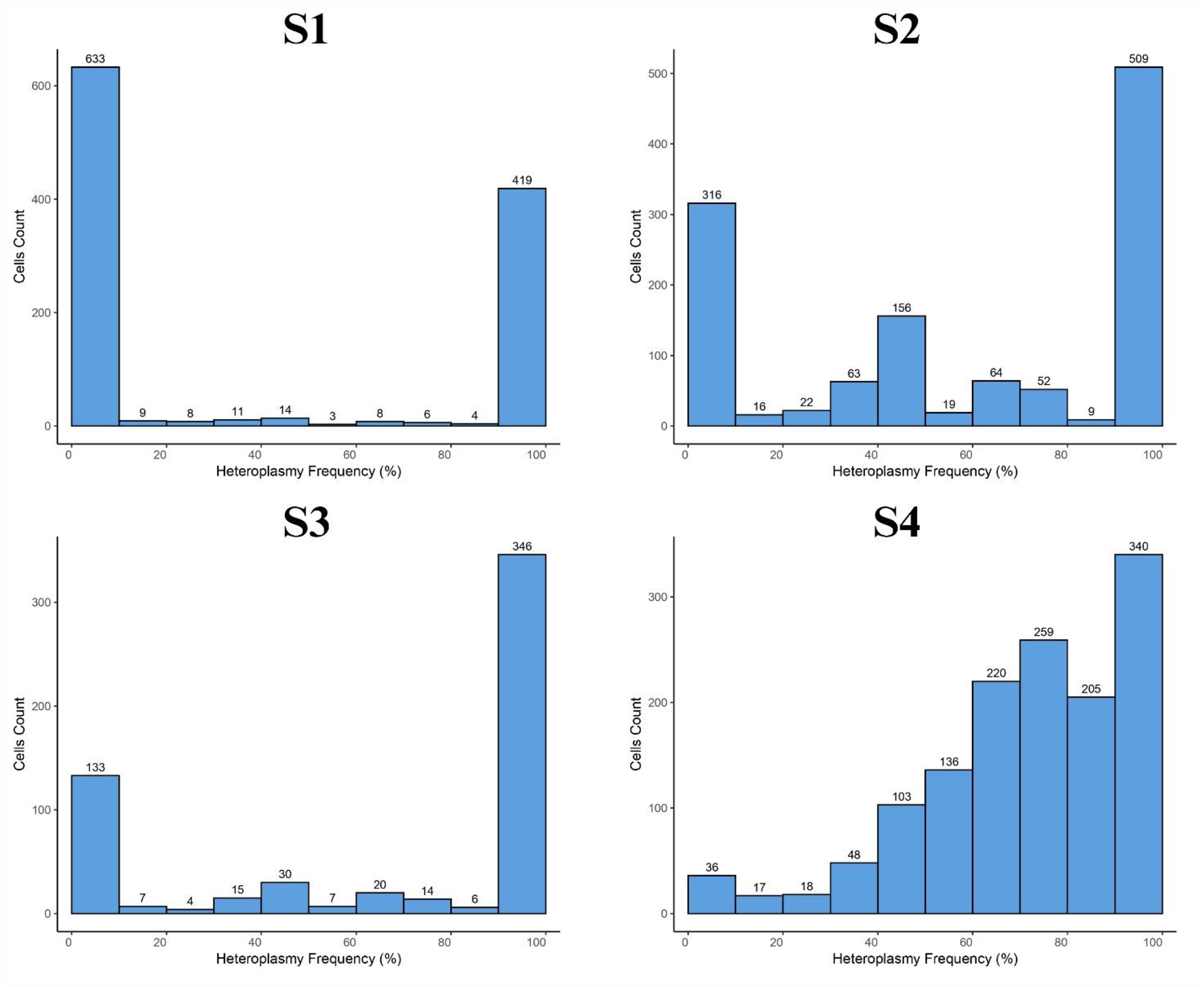
Distribution of per-cell heteroplasmy across four patient fibroblast samples. The y-axis represents the number of cells, and the x-axis shows heteroplasmy frequency in percentage with 10% histogram bins.

### 3.4 Transcriptomics effects of different RNA Heteroplasmy levels in the same subject/genetic background

To test whether high vs. low heteroplasmy defines discrete transcriptional states, on each cell line independently we exploited UMAP clustering, a technique used in data visualization to group cells with similar gene expression profiles into distinct clusters. Here we show sample S1 as the most representative example. Using a resolution of 0.3, all cells were stratified into four UMAP embedding clusters. Low- and High-heteroplasmy cells (in different colours in figure 3 on the right) remained fully interspersed across principal clusters, demonstrating that heteroplasmy level is not the defining factor in global transcriptomic clustering (Figure 3). The same pattern through intermixing between high and low heteroplasmy cells was observed in the other samples (Figure S2), although the sharpness of cluster definition varied with cell number.

**Figure 3.**
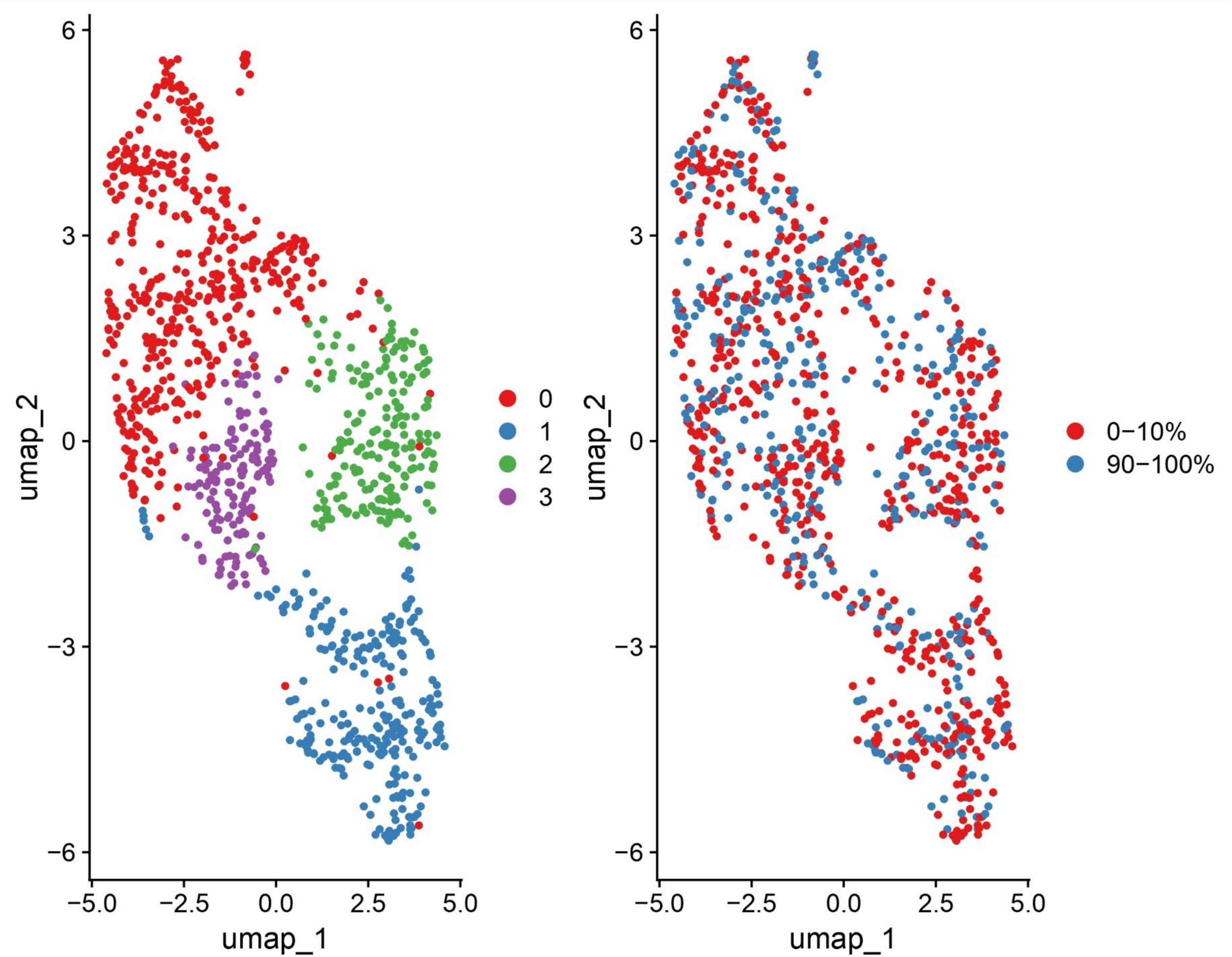
UMAP of sample S1 single-cell transcriptomes colored by heteroplasmy. Cells with high heteroplasmy levels (90–100%) are in red, while those with low heteroplasmy levels (0–10%) are in blue. The projection shows complete intermixing of both groups across clusters, indicating that heteroplasmy level does not drive major transcriptional variation.

To assess whether each cell line shows a distinct transcriptional response to high heteroplasmy (> 90%) at the individual-sample level, we performed sample-wise differential expression analysis and pathway enrichment analysis (Figure 4). Only genes expressed in at least 20% of cells were retained. Because no protein-coding gene survived a transcriptome-wide FDR correction, we defined candidate DEGs as those with a nominal LRT p-value (p(>Chisq) < 0.05) from the hurdle model. To further characterize modes of regulation, we extracted logFC, discrete (β_D), and continuous (β_C) hurdle model coefficients. For gene ontology (GO) pathway enrichment, we considered only terms with adjusted p-value < 0.05.

**Figure 4.**
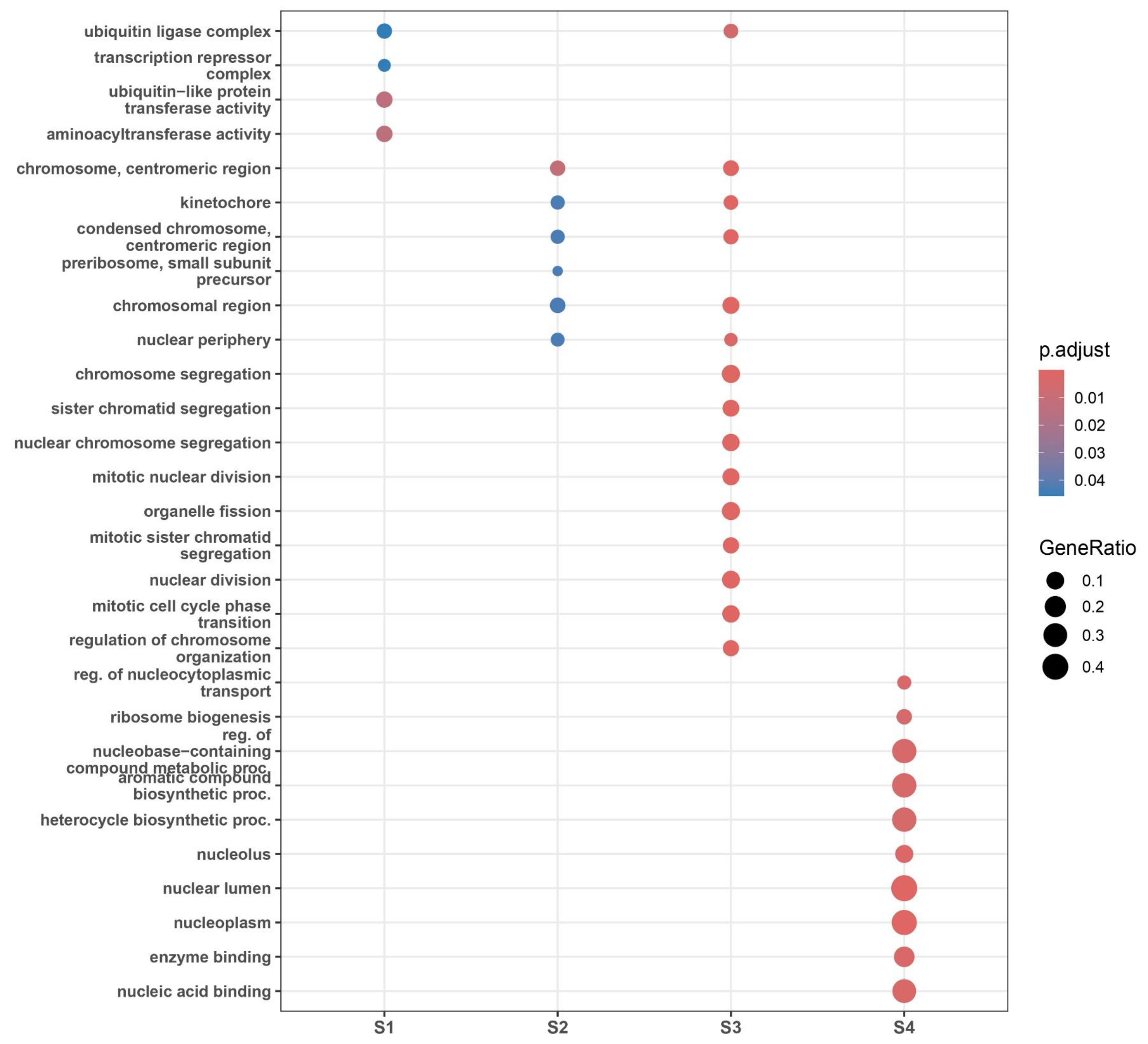
Comparative GO enrichment dot plot across four fibroblast samples (S1-S4) stratified by extreme heteroplasmy (> 90% vs. 0-10%) Each point represents a GO term significantly enriched (adjusted p-value < 0.05) in the DEGs of the corresponding sample. The y-axis lists the top 10 GO categories ranked by adjusted p-value, and the x-axis indicates aggregated samples. Point size reflects the number of DEGs contributing to the term, and color gradient represents the magnitude of p-adjust values (darker = more significant). This visualization highlights distinct transcriptional programs under high heteroplasmy: ubiquitin ligase activity in S1, chromosome-segregation structures in S2, mitotic and proteostasis pathways in S3, and RNA metabolism/post-transcriptional regulation in S4.

High heteroplasmy cells from S1 sample showed enrichment for ubiquitin-related and protein-quality control functions. By contrast, S2 sample exhibited enrichment almost exclusively in chromosome structural components, pointing to a potential delay or dysregulation of chromosomal segregation machinery under high heteroplasmy, which may impair faithful cell division. S3 cells showed broad engagement of mitotic checkpoints and proteostasis pathways, which suggests activation of cell-cycle arrest or slowing alongside enhanced proteasomal clearance. Finally, S4 displays yet another set of enriched categories: ribosome biogenesis, various RNA metabolic processes, and macromolecule biosynthesis. All four lines each exhibit a distinct transcriptional fingerprint under high heteroplasmy. These sample-specific responses underscore the heterogeneous cellular strategies possibly adopted to cope with mitochondrial genome imbalance.

To specifically assess the impact of extremely high heteroplasmy on the expression of genes with mitochondrial localization/function, we next performed a targeted enrichment of our nominal DEGs against the MitoCarta3.0 gene set, aiming to identify candidate-driven mitochondrial gene signatures characterizing each mutant background (Table S3).

In S1, we observed coordinated up-regulation of genes involved in Complex IV structure or assembly (COX7A1, SCO1) and mitochondrial translation (MRPS27, MTG2), together with an increased proportion of cells expressing COX10. By contrast, SLC25A20 and LIAS were down-regulated both in expression frequency and magnitude. In S2, a set of mitochondrial DNA maintenance and translation factors (e.g., POLRMT, LIG3, MRPL30, DNAJA3) and metabolic enzymes (e.g., ACAD10, IDH3A) genes showed changes almost entirely driven by the fraction of cells expressing each gene rather than by per-cell transcript abundance. Notably, the Complex I subunit gene MT-ND4L expression was detected in fewer cells under high heteroplasmy, whereas the Complex III assembly factor UQCC1 gene exhibited reduced transcript abundance per cell.

In S3, we observed combined up-regulation - in both the fraction of expressing cells and per-cell abundance - of the ATP synthase subunit MT-ATP8 (Complex V) and proteostasis/import genes (CLPP, ATAD3A, TOMM40), along with down-regulation of the replication factor POLG and antioxidant enzyme OXR1 in either expression frequency or magnitude. Finally, the S4 sample reveals a highly nuanced mitochondrial rewiring rather than a uniform shutdown. At the level of Complex I, we see a concerted down-regulation of core subunit genes - NDUFS1 and its assembly factor NDUFAF7 both lose expression in more cells and at lower levels - while NUBPL deviates from the trend with a subtle, continuous-only increase in transcript output among its remaining expressers. Complex II follows suit: both SDHA and SDHAF1 genes decline in frequency and magnitude, pointing to diminished succinate dehydrogenase activity. Downstream, we detect no classical subunit hits for Complex III, IV, or V.

Subsequent interrogation of our nominal DEGs against the MitoCarta3.0 MitoPathways repertoire revealed that, although all four fibroblast lines differentially express mitochondrial genes under extreme (> 90%) heteroplasmy, they do so with largely distinct repertoires. S2 shows the highest mitochondrial burden (8.2% of its 256 DEGs), closely followed by S4 (7.9% of its 391 DEGs), and then S1 and S3 at roughly 6% each of 332 and 533 DEGs, respectively. When we examine the overlap of these MitoCarta hits, the vast majority are unique to a single sample. Only a handful of genes are shared between pairs of lines - four genes overlap S3 and S4, two are common to S2 and S4, two to S1 and S4, and a single gene each is shared by S2–S3 and S1–S3. No mitochondrial gene is differentially expressed in more than three samples.

A complementary presence/absence map groups each gene by sample membership, clustering translation factors, OXPHOS subunits, and quality-control components, underscoring the sample-specific nature of the mitochondrial response to extreme heteroplasmy (Figure 5).

**Figure 5.**
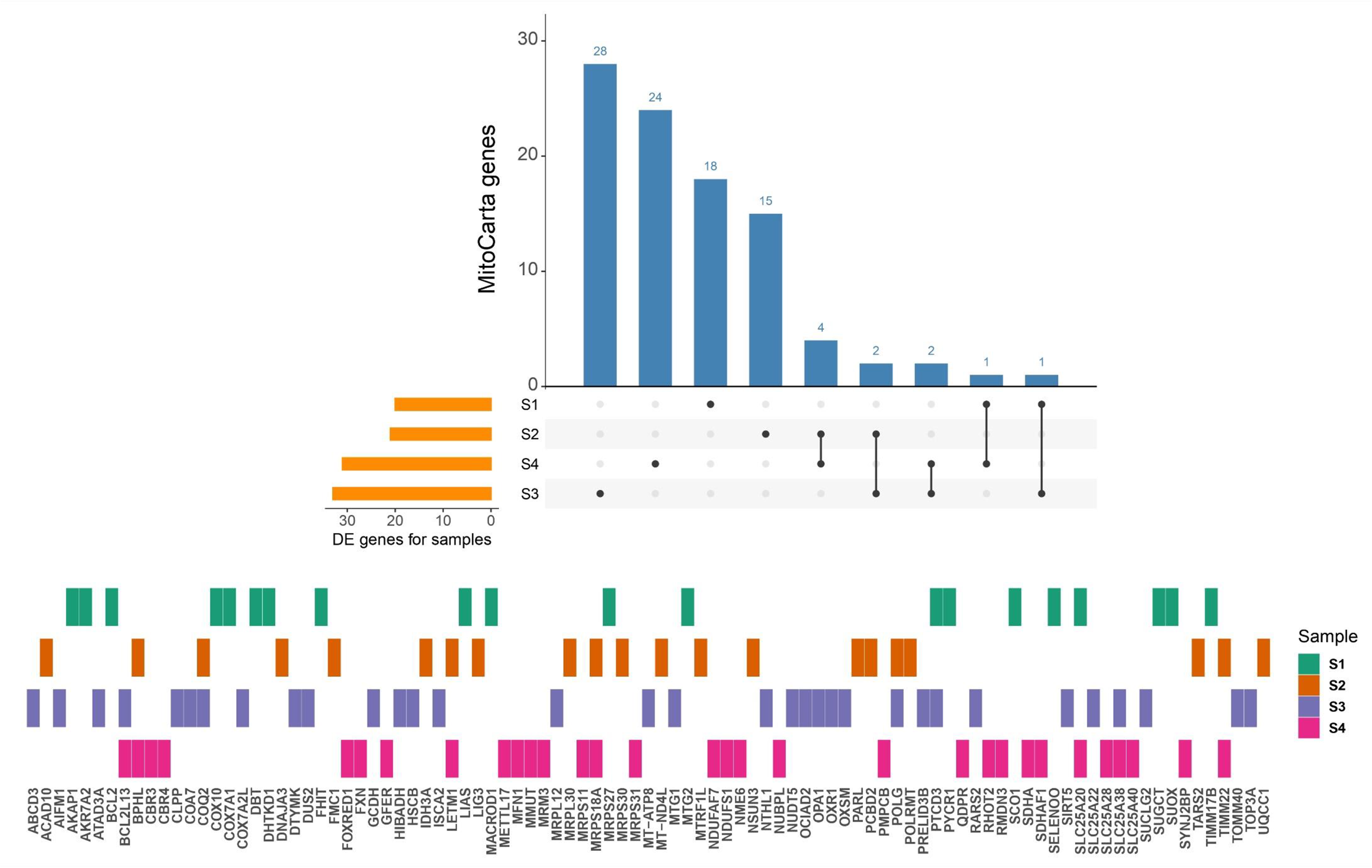
Intersection and presence/absence of MitoCarta3.0–annotated DEGs across four fibroblast samples under extreme heteroplasmy. In the UpSet plot (upper panel), the horizontal bars indicate the total number of mitochondrial DEGs per sample, and the vertical bars show the sizes of each intersection set. Dots and connecting lines beneath indicate which sample combinations contribute to each intersection. In Heat-tile (lower panel), each vertical tile represents a single MitoCarta DEG, colored by the samples in which it is differentially expressed. Tiles are grouped by sample membership, enabling visual identification of shared versus unique mitochondrial gene signatures. Colors: green = S1, orange = S2, purple = S3, magenta = S4.

### 3.5 Transcriptomics effects of high heteroplasmy for complex I mtDNA

Having characterized these sample-specific signatures, we next aggregated cells from all four samples with complex I subunit variants to uncover more subtle transcriptional effects of extreme heteroplasmy. We divided all cells from the 4 subjects S1-4 into high vs low heteroplasmy cells groups and performed single-cell differential expression analysis using the MAST hurdle model, with plate-level pooling and sample identity included as a fixed covariate.

In our analysis, we detected 502 genes whose expression differs between high- and low-heteroplasmy cells (LRT p < 0.05). Of these, only about 9% (42/502) are annotated in MitoCarta3.0 (Table S4).

This proportion is comparable to, or slightly higher than, the per-sample enrichments we observed previously (6-8%), indicating that pooling cells does not reveal a stronger core mitochondrial response, but rather recovers a modest “common” set of mitochondrial genes.

The mitochondrial hits encompass a mix of up- and down-regulation across OXPHOS complexes and their supporting machinery. In Complex I, for instance, some core subunit genes - like NDUFA9 – showed increased transcript levels, while other key components - like subunits MT-ND5 and NDUFV3 and assembly factor NDUFAF7 genes - are both expressed in fewer cells and at lower levels. A similar pattern emerges in Complex III: the assembly-factor gene UQCC1 is shut off in a subset of cells, even as other Complex III subunits remain unaffected, pointing to targeted remodeling of the bc1 machinery rather than global disassembly. Conversely, complex IV shows a differentiated strategy. The oxidase subunits SCO1 and SCO2 are up-regulated broadly, hinting at an effort to shore up electron-transfer capacity. In contrast, cytochrome c (CYCS) is silenced in many cells, and only a fraction retains expression of COX7A1, a key assembly scaffold, suggesting that Complex IV assembly is preserved in some cells, while compromised or absent in others. Finally, Complex V sees a modest, continuous-only drop in MT-ATP8 transcript levels within expressing cells, implying a nuanced throttling of ATP synthase output rather than a blanket shutdown.

Outside the core OXPHOS modules, expression of proteins related to mitochondrial translation is also rebalanced. For instance, MRPL55 and MRPL57 rise in cells that keep them on, indicating selective up-regulation of mitoribosome biogenesis. Meanwhile, CHCHD3 and MICOS10, encoding components of the inner-membrane organizing system, are pulled back, suggesting a contraction of membrane architecture support in some cells.

The integration of data across complex I deficient patients suggests the emergence of two distinct profiles: cells that sustain mitochondrial gene expression fine-tune specific OXPHOS subunits and translation machinery, whereas others quietly abandon key assembly factors. This heterogeneous rewiring underscores the complexity of cellular adaptation to extreme heteroplasmy.

Functional enrichments of total DEGs across all GO domains and KEGG pathways - each tested by Fisher’s exact test with Benjamini-Hochberg adjustment at p < 0.05 - highlight a widespread reprogramming across transport, proteostasis, and genome-stability pathways (Figure 6). At the cellular-component level, mitochondrial membrane organization features prominently, with recurrence of “integral component of mitochondrial inner membrane” and broader “mitochondrial inner membrane” terms, suggesting targeted remodeling of organelle architecture. In addition to these findings, when we turn to the KEGG database, the only significant hit is oxidative phosphorylation. Together, these shared enrichments highlight that, beyond transport and proteostasis, fine-tuning of energy-production machinery itself is a central shared feature of the high-heteroplasmy state

**Figure 6.**
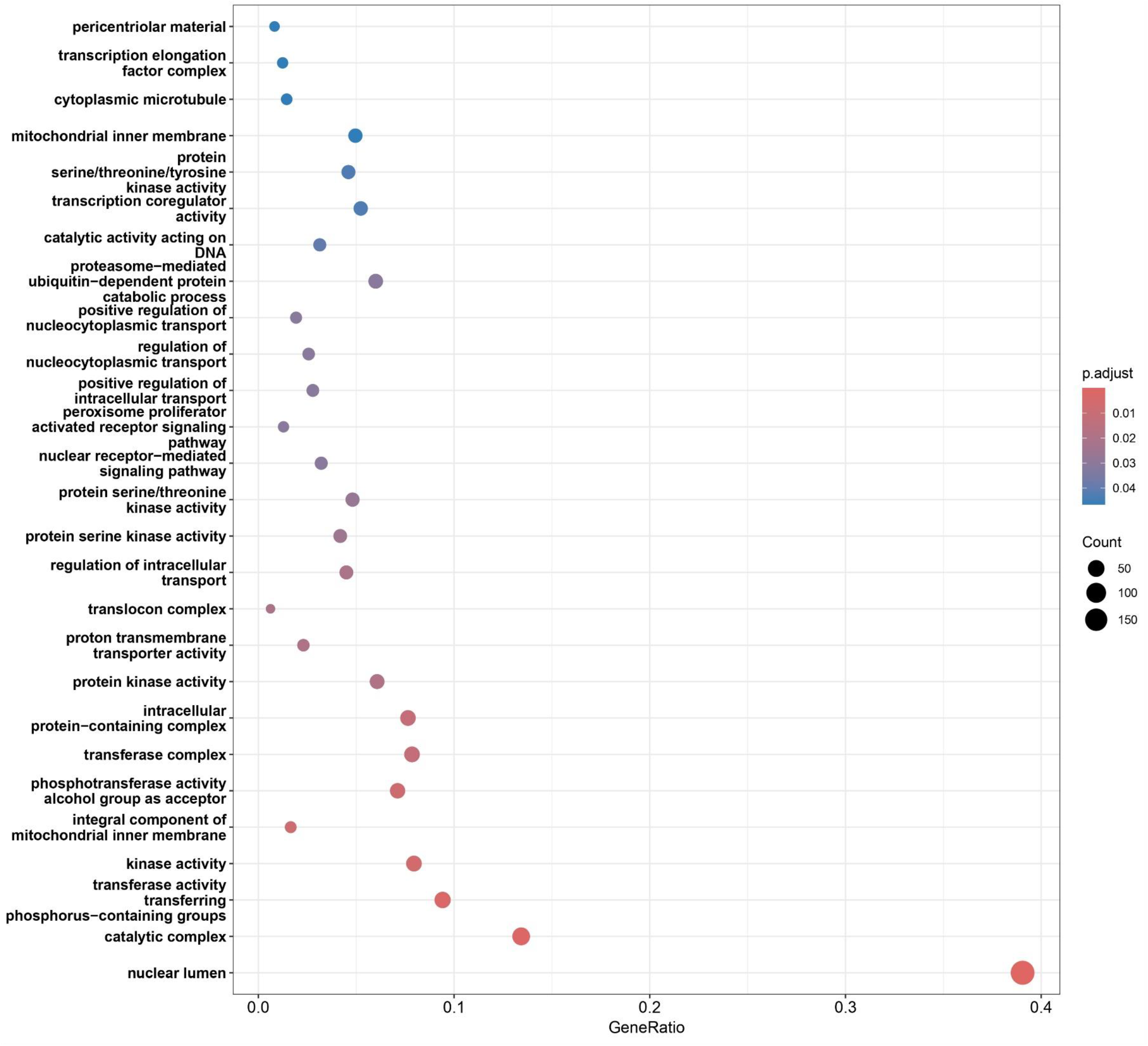
Combined top 10 GO enrichment summary for 502 genes differentially expressed between high- and low-heteroplasmy cells across all four fibroblast lines. GO dotplot: Each point represents a GO term colored by adjusted p-value (p.adjust) and sized by GeneRatio. Terms are split by ontology: Biological Process (BP) and Cellular Component (CC), and Molecular Function (MF). Notable BP enrichments include “regulation of intracellular transport,” “positive regulation of nucleocytoplasmic transport,” and “proteasome-mediated ubiquitin-dependent protein catabolic process”; CC highlights “mitochondrial inner membrane”, “integral component of mitochondrial inner membrane”; in MF, dominant activities include “protein serine/threonine kinase activity”.

## 4 Discussion

Using 10x Chromium 3′ scRNA-seq data, we demonstrate that mitochondrial RNA heteroplasmy can be estimated at single-cell resolution and that extreme heteroplasmy (>90%) is associated with both sample-specific transcriptional signatures and shared stress-response axes. scRNA heteroplasmy estimates generally agree with bulk RNA and DNA measurements, while per-cell analyses reveal marked heterogeneity within cell lines, including bimodal heteroplasmy distributions in three of four samples. Sample-wise and pooled analyses identify distinct mitochondrial and non-mitochondrial pathways engaged in response to high heteroplasmy.

As reported, our single-cell coverage analyses of mtDNA-encoded transcripts showed a strong 3′-end bias inherent to the 10x Chromium 3′ scRNA-seq chemistry, which limits the ability to uniformly capture long or poorly polyadenylated mitochondrial transcripts. Due to the low amount of starting material in single cells, protocols are optimized for efficiency over completeness. This leads to shallow coverage and a focus on quantifying gene expression rather than full-length transcript reconstruction. Most scRNA-seq protocols (e.g., 10x Chromium 3’ kit) are designed to capture polyadenylated mRNA using oligo(dT) primers. This means that reverse transcription starts at the poly(A) tail, which is located at the 3’-end of the transcript. As a result, only the 3’-end fragments of transcripts are reverse-transcribed and sequenced. In particular, MT-ND5 - the largest mtDNA-encoded gene at ~1.8 kb - shows the lowest and most uneven coverage, likely reflecting both its length and potential secondary structure hurdles to reverse transcriptase processivity, leading to drop-off and incomplete cDNA synthesis. Likewise, variable poly(A) tail length or stability in mitochondrial messages may further reduce capture efficiency. Longer transcripts like MT-ND5 may not be fully captured, especially if the poly(A) tail is distant from the coding region. Furthermore, mitochondrial transcripts, including MT-ND5, may have variable or inefficient polyadenylation, which affects their capture by oligo(dT) primers. These technical constraints make the reliability of per-cell heteroplasmy estimates dependent on the variant’s position within the transcript and its local sequence context. However, despite these biases, the obtained per-cell read counts for MT-ND4 and MT-ND5 were adequate to stratify cells into “low” (0–10%) and “high” (90–100%) heteroplasmy classes.

Despite discrepancies may occur between mtDNA heteroplasmy and the allele fractions inferred from transcript reads because variant-harbouring mitochondrial RNAs can be differentially unstable, degraded, or selectively transcribed/processed, our findings indicate that there are no active mechanisms against mutant mtDNA transcripts, at least for the investigated variants. The per-cell bimodal heteroplasmy distributions observed in S1–S3 - cells concentrated near 0-10% or 90-100% - may reflect intracellular segregation dynamics of mtDNA, selection for or against particular mitochondrial genotypes in culture, or differential turnover of mitochondrial genomes across subpopulations. The absence of significant differences in MT-ND4/MT-ND5 transcript abundance between High and Low groups argues against a simple model in which mutant transcripts are universally stabilized or over-transcribed across samples.

Our single-sample analyses revealed a remarkable diversity of mitochondrial responses to extreme heteroplasmy. Some lines (S1, S3) mount a “downstream compensation” by up-regulating Complex IV subunits and mitochondrial translation factors, presumably to boost electron flux around a deficient Complex I. Others (S2) appear to shift resources toward genome maintenance at the expense of respiratory-chain structure, as seen in selective suppression of Complex I/III components and up-regulation of mtDNA maintenance factors. This pattern suggests a strategic reduced priority of respiratory assembly in favor of preserving mitochondrial genome integrity. In the S4 line, we observed a targeted silencing of OXPHOS genes across multiple complexes. These contrasting strategies - from targeted downstream boosting (S1, S3), through genome-focused resource allocation (S2), to uniform shutdown and limited compensation (S4) - highlight how fibroblasts deploy sample-specific OXPHOS programs to contend with high burdens of Complex I variants.

When we aggregate all four sample lines into a single pseudo-bulk model, a coherent, cross-cutting stress response emerges. GO Biological Process enrichments center on mitotic checkpoints and chromosome dynamics, indicating that high heteroplasmy universally triggers tighter cell-cycle surveillance. Simultaneously, RNA-processing pathways are up-regulated, suggesting enhanced post-transcriptional quality control. Mitochondrial turnover also dominates: “autophagosome organization” and lysosomal components are consistently enriched, indicating escalated clearance of damaged organelles. KEGG hits about DNA replication and mismatch-repair pathways further underscore activation of genome-stability mechanisms under mitochondrial distress. Together, these shared axes - cell-cycle regulation, RNA quality control, selective organelle turnover, and genome-integrity enforcement - provide a framework for understanding possible common features of the high-heteroplasmy state, even as individual lines diverge in their mitochondrial-specific adaptations.

Building on these insights, we demonstrate a novel “virtual homoplasmic clone” approach: by stratifying cells from the same line into low- and high-heteroplasmy groups, we effectively create in silico cohorts of homoplasmic wild-type or mutant cells with the same genetic background that can be interrogated at scale. This strategy not only potentially enables robust differential expression analyses within individual patient lines but also provides a high-throughput framework for prioritizing mtDNA variants of uncertain significance for downstream validation and to probe genotype–phenotype relationships within a controlled, in-silico framework. Extending this strategy to larger cohorts of patient specimens (and possibly to research models such as organoids) and integrating it with multimodal single-cell data will help the dissection of pathogenic versus benign mitochondrial variants and to illuminate the cellular strategies that buffer or amplify mitochondrial dysfunction.

## 5 Conclusion

Our study shows that single-cell resolution of mtDNA heteroplasmy, coupled with a reproducible computational workflow, offers a powerful new window into cellular mitochondrial variability. At the aggregate level, RNA-derived heteroplasmy estimates agree with those obtained from DNA and bulk RNA; however, cell-by-cell analysis reveals a more nuanced reality: most cells tend to be at the extremes - close to wild-type or mutant homoplasmy - while only a small fraction show intermediate mutational loads. This pattern highlights how the same average heteroplasmy value can hide very different distributions at the cellular level, with important implications for understanding the penetrance and phenotypic variability of mitochondrial diseases.

Although high heteroplasmy is associated with transcriptional remodeling, these responses are largely sample-specific: each fibroblast line displays a distinct program of mitochondrial adaptation. Such diversity implies that the functional impact of a given mtDNA variant is strongly modulated by nuclear background and cellular context rather than governed by a single, uniform mechanism.

## Supporting information

Supplementary material including 4 tables and 2 figures

## Data Availability Statement

The data that support the findings of this study are available on request from the corresponding author.

## Ethics statement

Written informed consents for genetic analyses were obtained from the investigated subjects, according to the Declaration of Helsinki. Fibroblast lines were obtained from the “Cell line and DNA Bank of Genetic Movement Disorders and Mitochondrial Diseases” of the Telethon Network of Genetic Biobanks which stores biosamples with consent for research purposes (Ethical Committee CET4 Lombardia, 23/12/2024).

## Author Contributions

MB and AL implemented the bioinformatics analyses. IDM and AN performed scRNAseq experiments. EL provided genetic data and cell lines. MB, IDM, AN, AL and DG contributed to study set-up, evaluated and interpreted the data. MB and DG wrote the manuscript. All authors read, revised and approved the final manuscript.

## Acknowledgments

The “Cell line and DNA Bank of Genetic Movement Disorders and Mitochondrial Diseases” of the Telethon Network of Genetic Biobanks (GTB18001) supplied biological specimens.

## Supplementary material

Supplementary material associated with this article can be found in the online version. It includes 4 tables and 2 figures.

## References

Butler, A., Hoffman, P., Smibert, P., Papalexi, E., and Satija, R. (2018). Integrating single-cell transcriptomic data across different conditions, technologies, and species. Nat Biotechnol 36, 411–420. doi: 10.1038/nbt.4096

Cloonan, N., Forrest, A. R. R., Kolle, G., Gardiner, B. B. A., Faulkner, G. J., Brown, M. K., et al. (2008). Stem cell transcriptome profiling via massive-scale mRNA sequencing. Nat Methods 5, 613–619. doi: 10.1038/nmeth.1223

Danielski, K. (2023). Guidance on Processing the 10x Genomics Single Cell Gene Expression Assay. Methods Mol Biol 2584, 1–28. doi: 10.1007/978-1-0716-2756-3_1

Finak, G., McDavid, A., Yajima, M., Deng, J., Gersuk, V., Shalek, A. K., et al. (2015). MAST: a flexible statistical framework for assessing transcriptional changes and characterizing heterogeneity in single-cell RNA sequencing data. Genome Biology 16, 278. doi: 10.1186/s13059-015-0844-5

Lareau, C. A., Dubois, S. M., Buquicchio, F. A., Hsieh, Y.-H., Garg, K., Kautz, P., et al. (2023). Single-cell multi-omics of mitochondrial DNA disorders reveals dynamics of purifying selection across human immune cells. Nat Genet 55, 1198–1209. doi: 10.1038/s41588-023-01433-8

Ludwig, L. S., Lareau, C. A., Ulirsch, J. C., Christian, E., Muus, C., Li, L. H., et al. (2019). Lineage Tracing in Humans Enabled by Mitochondrial Mutations and Single-Cell Genomics. Cell 176, 1325–1339.e22. doi: 10.1016/j.cell.2019.01.022

Marshall, A. S., and Jones, N. S. (2021). Discovering Cellular Mitochondrial Heteroplasmy Heterogeneity with Single Cell RNA and ATAC Sequencing. Biology (Basel) 10, 503. doi: 10.3390/biology10060503

Miller, T. E., Lareau, C. A., Verga, J. A., DePasquale, E. A. K., Liu, V., Ssozi, D., et al. (2022). Mitochondrial variant enrichment from high-throughput single-cell RNA-seq resolves clonal populations. Nat Biotechnol 40, 1030–1034. doi: 10.1038/s41587-022-01210-8

Nakagawa, H., Shima, Y., Sasagawa, Y., and Nikaido, I. (2025). MitoDelta: identifying mitochondrial DNA deletions at cell-type resolution from single-cell RNA sequencing data. 2025.06.10.658806. doi: 10.1101/2025.06.10.658806

Nissanka, N., and Moraes, C. T. (2020). Mitochondrial DNA heteroplasmy in disease and targeted nuclease-based therapeutic approaches. EMBO reports 21, e49612. doi: 10.15252/embr.201949612

Nitsch, L., Lareau, C. A., and Ludwig, L. S. (2024). Mitochondrial genetics through the lens of single-cell multi-omics. Nat Genet 56, 1355–1365. doi: 10.1038/s41588-024-01794-8

Parakatselaki, M.-E., and Ladoukakis, E. D. (2021). mtDNA Heteroplasmy: Origin, Detection, Significance, and Evolutionary Consequences. Life 11, 633. doi: 10.3390/life11070633

Payne, B. A. I., Wilson, I. J., Yu-Wai-Man, P., Coxhead, J., Deehan, D., Horvath, R., et al. (2013). Universal heteroplasmy of human mitochondrial DNA. Hum Mol Genet 22, 384–390. doi: 10.1093/hmg/dds435

Stewart, J. B., and Chinnery, P. F. (2021). Extreme heterogeneity of human mitochondrial DNA from organelles to populations. Nat Rev Genet 22, 106–118. doi: 10.1038/s41576-020-00284-x

Wang, M., Deng, W., Samuels, D. C., Zhao, Z., and Simon, L. M. (2023). MitoTrace: A Computational Framework for Analyzing Mitochondrial Variation in Single-Cell RNA Sequencing Data. Genes 14, 1222. doi: 10.3390/genes14061222

