## Supplementary material including 4 tables and 2 figures for "Exploring Heteroplasmic Variants in mtDNA: Insights from Single-Cell Transcriptomics"

### Supplemetary material

Supplementary material includes 4 tables and 2 figures.

**Supplementary Table S1. Key statistics (Wilcoxon test, FDR-adjusted p > 0.4 for all comparisons) in raw counts**

| sample | gene | med_90_100 | med_0_10 | $log_2FC$[1] | p_value | p_adj |
|---|---|---|---|---|---|---|
| S1 | *MT-ND4* | 136 | 138 | -0.02106 | 0.151703 | 0.466904 |
| S1 | *MT-ND5* | 22 | 23 | -0.06413 | 0.117216 | 0.466904 |
| S2 | *MT-ND4* | 145.5 | 147.5 | -0.0197 | 0.625216 | 0.725321 |
| S2 | *MT-ND5* | 27 | 28 | -0.05247 | 0.422222 | 0.675556 |
| S3 | *MT-ND4* | 137.5 | 135 | 0.026472 | 0.370624 | 0.675556 |
| S3 | *MT-ND5* | 12 | 13 | -0.11548 | 0.175089 | 0.466904 |
| S4 | *MT-ND4* | 169 | 174.5 | -0.0462 | 0.854571 | 0.854571 |
| S4 | *MT-ND5* | 22 | 24.5 | -0.15528 | 0.634656 | 0.725321 |

[1] $log_2FC$ calculated as $log_2(median_{90-100}\ \% / median_{0-10}\ \%)$.

**Supplementary Table S2. Key statistics (Wilcoxon test, FDR-adjusted p > 0.4 for all comparisons) in normalized data counts.**

| sample | gene | med_90_100 | med_0_10 | $log_2FC$[1] | p_value | p_adj |
|---|---|---|---|---|---|---|
| S1 | *MT-ND4* | 3.53534 | 3.510103 | 1.951595 | 0.533223 | 0.73173 |
| S1 | *MT-ND5* | 1.830181 | 1.867826 | 2.030465 | 0.172276 | 0.73173 |
| S2 | *MT-ND4* | 3.77063 | 3.768152 | 1.970173 | 0.593752 | 0.73173 |
| S2 | *MT-ND5* | 2.151444 | 2.198687 | 1.892807 | 0.317676 | 0.73173 |
| S3 | *MT-ND4* | 3.437477 | 3.423126 | 1.936264 | 0.927116 | 0.927116 |
| S3 | *MT-ND5* | 1.284314 | 1.35311 | 2.94379 | 0.210773 | 0.73173 |
| S4 | *MT-ND4* | 3.306982 | 3.203331 | 1.968943 | 0.49022 | 0.73173 |
| S4 | *MT-ND5* | 1.539263 | 1.530917 | 2.505327 | 0.640264 | 0.73173 |

[1] $log_2FC$ calculated as $log_2(median_{90-100}\ \% / median_{0-10}\ \%)$.

**Supplementary Table S3. MitoCarta3.0–annotated differentially expressed genes in each sample under extreme heteroplasmy (> 90 % vs. 0–10 %).**

| **S1 sample** | | | | | | | | | | |
|---|---|---|---|---|---|---|---|---|---|---|
| **primerid** | **Pr(>Chisq)** | **coef** | **ci.hi** | **ci.lo** | **fdr** | **fold_change[1]** | **log$_2$FC[2]** | **β_D** | **β_C** | **category** |
| *AKAP1* | 0.0297 | 0.1036 | 0.1810 | 0.0261 | 0.9978 | 1.1091 | 0.1494 | 0.3393 | 0.0329 | up |
| *AKR7A2* | 0.0100 | 0.2068 | 0.3422 | 0.0714 | 0.9978 | 1.2297 | 0.2983 | 0.3814 | 0.1497 | up |
| *BCL2* | 0.0238 | 0.0072 | 0.1021 | -0.0877 | 0.9978 | 1.0072 | 0.0104 | 0.1798 | -0.1555 | D_driver |
| *COX10* | 0.0015 | 0.1008 | 0.1794 | 0.0223 | 0.9978 | 1.1061 | 0.1455 | 0.4258 | -0.0548 | D_driver |
| *COX7A1* | 0.0042 | 0.1960 | 0.3348 | 0.0572 | 0.9978 | 1.2165 | 0.2828 | 0.4398 | 0.0063 | up |
| *DBT* | 0.0319 | -0.0194 | 0.0500 | -0.0888 | 0.9978 | 0.9808 | -0.0280 | -0.1788 | 0.0810 | D_driver |
| *DHTKD1* | 0.0373 | 0.0570 | 0.1348 | -0.0208 | 0.9978 | 1.0587 | 0.0822 | 0.2748 | -0.0558 | D_driver |
| *FHIT* | 0.0121 | -0.0543 | 0.0524 | -0.1611 | 0.9978 | 0.9471 | -0.0784 | 0.0733 | -0.1799 | C_driver |
| *LIAS* | 0.0413 | -0.0831 | -0.0151 | -0.1511 | 0.9978 | 0.9203 | -0.1199 | -0.3545 | 0.0045 | D_driver |
| *MACROD1* | 0.0403 | 0.0434 | 0.1470 | -0.0602 | 0.9978 | 1.0443 | 0.0626 | -0.0704 | 0.1019 | C_driver |
| *MRPS27* | 0.0281 | 0.0582 | 0.1613 | -0.0450 | 0.9978 | 1.0599 | 0.0839 | 0.3075 | -0.0607 | D_driver |
| *MTG2* | 0.0098 | 0.1124 | 0.2011 | 0.0236 | 0.9978 | 1.1189 | 0.1621 | 0.2030 | 0.0932 | up |
| *PTCD3* | 0.0050 | 0.0049 | 0.1042 | -0.0944 | 0.9978 | 1.0049 | 0.0071 | -0.2264 | 0.1086 | C_driver |
| *PYCR1* | 0.0496 | -0.0095 | 0.0716 | -0.0907 | 0.9978 | 0.9905 | -0.0138 | 0.0936 | -0.1004 | C_driver |
| *SCO1* | 0.0207 | 0.1395 | 0.2383 | 0.0406 | 0.9978 | 1.1496 | 0.2012 | 0.3453 | 0.0424 | up |
| *SELENOO* | 0.0232 | -0.0219 | 0.0683 | -0.1120 | 0.9978 | 0.9783 | -0.0316 | 0.1020 | -0.0934 | C_driver |
| *SLC25A20* | 0.0091 | -0.0932 | -0.0259 | -0.1605 | 0.9978 | 0.9110 | -0.1345 | -0.2914 | -0.0727 | down |
| *SUGCT* | 0.0384 | -0.1397 | -0.0287 | -0.2507 | 0.9978 | 0.8696 | -0.2015 | -0.2163 | -0.1130 | down |
| *SUOX* | 0.0377 | 0.0755 | 0.1355 | 0.0155 | 0.9978 | 1.0784 | 0.1090 | 0.3920 | 0.0065 | up |
| *TIMM17B* | 0.0204 | -0.1076 | 0.0053 | -0.2204 | 0.9978 | 0.8980 | -0.1552 | -0.3776 | 0.0297 | D_driver |

| **S2 sample** | | | | | | | | | | |
|---|---|---|---|---|---|---|---|---|---|---|
| **primerid** | **Pr(>Chisq)** | **coef** | **ci.hi** | **ci.lo** | **fdr** | **fold_change[1]** | **log2FC[2]** | **β_D** | **β_C** | **category** |
| *ACAD10* | 0.0455 | 0.0403 | 0.1045 | -0.0239 | 0.9999 | 1.0411 | 0.0581 | 0.3012 | -0.0683 | D_driver |
| *BPHL* | 0.0097 | 0.0824 | 0.1626 | 0.0022 | 0.9999 | 1.0859 | 0.1189 | 0.4132 | -0.0675 | D_driver |
| *COQ2* | 0.0087 | -0.0843 | 0.0244 | -0.1931 | 0.9999 | 0.9191 | -0.1217 | -0.3746 | 0.0809 | D_driver |
| *DNAJA3* | 0.0066 | 0.0398 | 0.1283 | -0.0487 | 0.9999 | 1.0406 | 0.0574 | 0.2836 | -0.1141 | D_driver |
| *FMC1* | 0.0109 | -0.0370 | 0.0469 | -0.1209 | 0.9999 | 0.9637 | -0.0533 | -0.2613 | 0.0935 | D_driver |
| *IDH3A* | 0.0284 | -0.0389 | 0.0599 | -0.1378 | 0.9999 | 0.9618 | -0.0562 | -0.2557 | 0.0916 | D_driver |
| *LETM1* | 0.0446 | -0.0028 | 0.0903 | -0.0959 | 0.9999 | 0.9972 | -0.0040 | -0.1425 | 0.0991 | D_driver |
| *LIG3* | 0.0115 | 0.0932 | 0.1667 | 0.0197 | 0.9999 | 1.0977 | 0.1344 | 0.4614 | -0.0388 | D_driver |
| *MRPL30* | 0.0248 | 0.0277 | 0.1141 | -0.0588 | 0.9999 | 1.0281 | 0.0399 | 0.2250 | -0.0991 | D_driver |
| *MRPS18A* | 0.0118 | -0.1383 | -0.0197 | -0.2570 | 0.9999 | 0.8708 | -0.1996 | -0.4367 | 0.0236 | D_driver |
| *MRPS30* | 0.0466 | -0.0377 | 0.0632 | -0.1385 | 0.9999 | 0.9630 | -0.0543 | -0.2379 | 0.0797 | D_driver |
| *MT-ND4L* | 0.0330 | -0.0149 | 0.0984 | -0.1282 | 0.9999 | 0.9852 | -0.0216 | -1.7373 | 0.0367 | D_driver |
| *MTRF1L* | 0.0058 | 0.1114 | 0.2112 | 0.0117 | 0.9999 | 1.1179 | 0.1608 | 0.1387 | 0.1358 | up |
| *NSUN3* | 0.0192 | -0.0556 | 0.0178 | -0.1290 | 0.9999 | 0.9459 | -0.0802 | -0.1355 | -0.1123 | down |
| *PARL* | 0.0453 | -0.0522 | 0.0432 | -0.1476 | 0.9999 | 0.9492 | -0.0753 | -0.2674 | 0.0702 | D_driver |
| *PCBD2* | 0.0457 | 0.1351 | 0.2450 | 0.0252 | 0.9999 | 1.1446 | 0.1949 | 0.3495 | 0.0265 | up |
| *POLG* | 0.0474 | -0.0581 | 0.0383 | -0.1545 | 0.9999 | 0.9435 | -0.0838 | -0.2840 | 0.0712 | D_driver |
| *POLRMT* | 0.0032 | 0.1025 | 0.1811 | 0.0238 | 0.9999 | 1.1079 | 0.1479 | 0.5069 | -0.0664 | D_driver |

| | | | | | | | | | | |
|---|---|---|---|---|---|---|---|---|---|---|
| *TARS2* | 0.0267 | -0.0613 | 0.0165 | -0.1391 | 0.9999 | 0.9405 | -0.0884 | -0.3305 | 0.0624 | D_driver |
| *TIMM22* | 0.0411 | 0.0273 | 0.1437 | -0.0890 | 0.9999 | 1.0277 | 0.0394 | -0.1321 | 0.1124 | C_driver |
| *UQCC1* | 0.0051 | -0.0500 | 0.0532 | -0.1531 | 0.9999 | 0.9513 | -0.0721 | 0.0529 | -0.1492 | C_driver |

| **S3 sample** | | | | | | | | | | |
|---|---|---|---|---|---|---|---|---|---|---|
| **primerid** | **Pr(>Chisq)** | **coef** | **ci.hi** | **ci.lo** | **fdr** | **fold_change[1]** | **log2FC[2]** | **β_D** | **β_C** | **category** |
| *ABCD3* | 0.0356 | -0.1285 | 0.0404 | -0.2975 | 0.9025 | 0.8794 | -0.1854 | -0.0605 | -0.1617 | down |
| *AIFM1* | 0.0288 | 0.1366 | 0.2956 | -0.0224 | 0.8965 | 1.1464 | 0.1971 | 0.0635 | 0.1741 | up |
| *ATAD3A* | 0.0112 | 0.2397 | 0.4014 | 0.0780 | 0.8663 | 1.2709 | 0.3458 | 0.6220 | 0.0598 | up |
| *BCL2L13* | 0.0349 | 0.2164 | 0.3794 | 0.0534 | 0.9025 | 1.2416 | 0.3121 | 0.4376 | 0.1092 | up |
| *CLPP* | 0.0279 | 0.2283 | 0.4303 | 0.0263 | 0.8934 | 1.2564 | 0.3293 | 0.8299 | 0.0577 | up |
| *COA7* | 0.0241 | 0.1244 | 0.2559 | -0.0072 | 0.8934 | 1.1324 | 0.1794 | 0.5235 | -0.0750 | D_driver |
| *COQ2* | 0.0481 | 0.0982 | 0.2659 | -0.0696 | 0.9025 | 1.1032 | 0.1416 | 0.4232 | -0.1000 | D_driver |
| *COX7A2L* | 0.0260 | -0.0187 | 0.2288 | -0.2662 | 0.8934 | 0.9815 | -0.0270 | 0.8782 | -0.1770 | C_driver |
| *DTYMK* | 0.0379 | 0.3186 | 0.5820 | 0.0553 | 0.9025 | 1.3753 | 0.4597 | 0.6290 | 0.1328 | up |
| *DUS2* | 0.0435 | -0.1062 | 0.0095 | -0.2219 | 0.9025 | 0.8992 | -0.1532 | -0.4953 | 0.0567 | D_driver |
| *GCDH* | 0.0113 | 0.1882 | 0.3067 | 0.0697 | 0.8663 | 1.2070 | 0.2715 | 0.5581 | 0.0982 | up |
| *HIBADH* | 0.0304 | 0.0826 | 0.2493 | -0.0840 | 0.9024 | 1.0861 | 0.1192 | 0.4467 | -0.1055 | D_driver |
| *HSCB* | 0.0247 | -0.1833 | -0.0467 | -0.3198 | 0.8934 | 0.8326 | -0.2644 | -0.4823 | -0.0722 | down |
| *ISCA2* | 0.0344 | 0.2135 | 0.3822 | 0.0447 | 0.9025 | 1.2380 | 0.3080 | 0.3585 | 0.1472 | up |
| *MRPL12* | 0.0197 | 0.2203 | 0.4159 | 0.0247 | 0.8934 | 1.2464 | 0.3178 | 0.0785 | 0.2269 | up |
| *MT-ATP8* | 0.0346 | 0.0019 | 0.1920 | -0.1882 | 0.9025 | 1.0019 | 0.0028 | 0.4119 | -0.1496 | D_driver |
| *MTG1* | 0.0497 | 0.1795 | 0.3359 | 0.0231 | 0.9025 | 1.1966 | 0.2590 | 0.2880 | 0.1353 | up |
| *NTHL1* | 0.0367 | 0.0528 | 0.2121 | -0.1066 | 0.9025 | 1.0542 | 0.0761 | 0.3427 | -0.1329 | D_driver |
| *NUDT5* | 0.0451 | 0.1993 | 0.3910 | 0.0076 | 0.9025 | 1.2205 | 0.2875 | 0.9393 | 0.0598 | up |
| *OCIAD2* | 0.0189 | 0.1453 | 0.3512 | -0.0606 | 0.8934 | 1.1564 | 0.2097 | 0.5217 | -0.2081 | D_driver |
| *OPA1* | 0.0312 | 0.0639 | 0.2454 | -0.1176 | 0.9025 | 1.0660 | 0.0922 | 0.5643 | -0.1058 | D_driver |
| *OXR1* | 0.0457 | -0.0326 | 0.1807 | -0.2459 | 0.9025 | 0.9679 | -0.0470 | 0.7170 | -0.1550 | C_driver |
| *OXSM* | 0.0212 | -0.1259 | -0.0047 | -0.2472 | 0.8934 | 0.8817 | -0.1817 | -0.5471 | 0.0579 | D_driver |
| *POLG* | 0.0055 | -0.0562 | 0.0856 | -0.1980 | 0.8646 | 0.9454 | -0.0811 | 0.0857 | -0.1810 | C_driver |
| *PRELID3B* | 0.0368 | 0.0192 | 0.1952 | -0.1568 | 0.9025 | 1.0194 | 0.0277 | -0.5528 | 0.1443 | C_driver |
| *PTCD3* | 0.0304 | 0.1533 | 0.3265 | -0.0198 | 0.9024 | 1.1657 | 0.2212 | 0.6076 | -0.0393 | D_driver |
| *RARS2* | 0.0388 | 0.0032 | 0.1799 | -0.1736 | 0.9025 | 1.0032 | 0.0046 | 0.5194 | -0.1270 | D_driver |
| *SIRT5* | 0.0319 | -0.1232 | -0.0014 | -0.2450 | 0.9025 | 0.8841 | -0.1777 | -0.3243 | -0.1170 | down |
| *SLC25A22* | 0.0434 | 0.0337 | 0.1587 | -0.0912 | 0.9025 | 1.0343 | 0.0487 | 0.2987 | -0.1431 | D_driver |
| *SLC25A38* | 0.0499 | 0.1323 | 0.2718 | -0.0073 | 0.9025 | 1.1414 | 0.1908 | 0.4988 | -0.0550 | D_driver |
| *SUCLG2* | 0.0229 | 0.0647 | 0.2699 | -0.1404 | 0.8934 | 1.0669 | 0.0934 | 0.7952 | -0.1108 | D_driver |
| *TOMM40* | 0.0263 | 0.2453 | 0.4489 | 0.0418 | 0.8934 | 1.2780 | 0.3539 | 0.6622 | 0.0491 | up |
| *TOP3A* | 0.0151 | 0.1598 | 0.2716 | 0.0480 | 0.8785 | 1.1733 | 0.2305 | 0.4770 | 0.1176 | up |

| **S4 sample** | | | | | | | | | | |
|---|---|---|---|---|---|---|---|---|---|---|
| **primerid** | **Pr(>Chisq)** | **coef** | **ci.hi** | **ci.lo** | **fdr** | **fold_change[1]** | **log2FC[2]** | **β_D** | **β_C** | **category** |
| *BCL2L13* | 0.0404 | -0.2169 | 0.1214 | -0.5551 | 0.9917 | 0.8050 | -0.3129 | -0.0047 | -0.3133 | down |
| *BPHL* | 0.0447 | -0.0178 | 0.2097 | -0.2452 | 0.9917 | 0.9824 | -0.0256 | 0.2387 | -0.2622 | C_driver |
| *CBR3* | 0.0171 | -0.0952 | 0.2747 | -0.4650 | 0.9917 | 0.9092 | -0.1373 | 0.2842 | -0.4284 | C_driver |
| *CBR4* | 0.0262 | -0.3019 | 0.0088 | -0.6125 | 0.9917 | 0.7394 | -0.4355 | -0.3574 | -0.2744 | down |
| *FOXRED1* | 0.0252 | -0.1432 | 0.1118 | -0.3983 | 0.9917 | 0.8665 | -0.2067 | -0.1492 | -0.2778 | down |

| | | | | | | | | | |
|---|---|---|---|---|---|---|---|---|---|
| *FXN* | 0.0464 | -0.3518 | -0.0658 | -0.6379 | 0.9917 | 0.7034 | -0.5076 | -0.7376 | -0.1709 | down |
| *GFER* | 0.0081 | 0.0277 | 0.4097 | -0.3544 | 0.9909 | 1.0280 | 0.0399 | 0.6982 | -0.3629 | D_driver |
| *LETM1* | 0.0090 | 0.3781 | 0.6164 | 0.1397 | 0.9909 | 1.4594 | 0.5454 | 0.6584 | 0.3125 | up |
| *METTL17* | 0.0024 | -0.4097 | -0.1740 | -0.6454 | 0.9909 | 0.6639 | -0.5910 | -1.2847 | -0.0601 | down |
| *MFN1* | 0.0203 | 0.0186 | 0.2719 | -0.2347 | 0.9917 | 1.0188 | 0.0269 | -0.6050 | 0.2588 | C_driver |
| *MMUT* | 0.0142 | -0.3216 | -0.0780 | -0.5652 | 0.9909 | 0.7250 | -0.4640 | -1.1930 | 0.0223 | D_driver |
| *MRM3* | 0.0144 | -0.2702 | 0.0847 | -0.6250 | 0.9909 | 0.7632 | -0.3898 | -0.1160 | -0.4338 | down |
| *MRPS11* | 0.0451 | -0.4436 | -0.0715 | -0.8156 | 0.9917 | 0.6418 | -0.6399 | -0.5562 | -0.3615 | down |
| *MRPS18A* | 0.0147 | -0.3015 | 0.0742 | -0.6771 | 0.9909 | 0.7397 | -0.4349 | 0.1040 | -0.4049 | C_driver |
| *MRPS31* | 0.0207 | -0.4857 | -0.1585 | -0.8129 | 0.9917 | 0.6153 | -0.7007 | -1.2815 | -0.2425 | down |
| *NDUFAF7* | 0.0406 | -0.3194 | -0.0614 | -0.5774 | 0.9917 | 0.7266 | -0.4608 | -0.8312 | -0.1080 | down |
| *NDUFS1* | 0.0366 | -0.3959 | -0.1171 | -0.6747 | 0.9917 | 0.6731 | -0.5712 | -1.3419 | -0.2146 | down |
| *NME6* | 0.0004 | -0.3651 | -0.1095 | -0.6207 | 0.5579 | 0.6941 | -0.5268 | -0.9051 | -0.2507 | down |
| *NUBPL* | 0.0261 | -0.0089 | 0.1946 | -0.2125 | 0.9917 | 0.9911 | -0.0129 | 0.2902 | -0.3299 | C_driver |
| *PMPCB* | 0.0403 | -0.4159 | -0.0893 | -0.7424 | 0.9917 | 0.6598 | -0.6000 | -0.7036 | -0.3166 | down |
| *QDPR* | 0.0052 | 0.0568 | 0.5389 | -0.4253 | 0.9909 | 1.0584 | 0.0819 | 1.0651 | -0.4038 | D_driver |
| *RHOT2* | 0.0244 | -0.3251 | -0.0235 | -0.6268 | 0.9917 | 0.7224 | -0.4691 | -0.4639 | -0.2638 | down |
| *RMDN3* | 0.0105 | 0.1285 | 0.4209 | -0.1639 | 0.9909 | 1.1372 | 0.1854 | 0.7701 | -0.2793 | D_driver |
| *SDHA* | 0.0406 | -0.4190 | -0.1054 | -0.7326 | 0.9917 | 0.6577 | -0.6045 | -0.9334 | -0.2434 | down |
| *SDHAF1* | 0.0334 | -0.3545 | -0.0878 | -0.6211 | 0.9917 | 0.7016 | -0.5114 | -0.8946 | -0.1084 | down |
| *SLC25A20* | 0.0399 | -0.0119 | 0.2582 | -0.2820 | 0.9917 | 0.9882 | -0.0171 | 0.3000 | -0.2501 | C_driver |
| *SLC25A28* | 0.0186 | -0.3132 | -0.0159 | -0.6104 | 0.9917 | 0.7311 | -0.4518 | -0.4669 | -0.2642 | down |
| *SLC25A38* | 0.0107 | -0.2195 | 0.1537 | -0.5928 | 0.9909 | 0.8029 | -0.3167 | 0.0643 | -0.5162 | C_driver |
| *SLC25A40* | 0.0232 | -0.1021 | 0.1286 | -0.3328 | 0.9917 | 0.9029 | -0.1473 | -0.0845 | -0.2974 | down |
| *SYNJ2BP* | 0.0354 | -0.2394 | 0.0875 | -0.5663 | 0.9917 | 0.7871 | -0.3453 | -0.1377 | -0.2957 | down |
| *TIMM22* | 0.0157 | -0.4145 | -0.0264 | -0.8026 | 0.9917 | 0.6607 | -0.5980 | -0.2152 | -0.4212 | down |

[1] fold-change = (exp(coef)). [2]$\log_2$FC: base-2 log-fold-change (coef / log(2)).
Each row lists a gene (**primerid**) passing the nominal LRT p-value cutoff (Pr(>Chisq) < 0.05). **Pr(>Chisq)**: likelihood-ratio test p-value for the cond90-100% contrast; **coef** (log fold-change) and its 95 % confidence interval (**ci.hi, ci.lo**), representing the overall log-scale difference in expression; **fdr**: adjusted p-value (Benjamini–Hochberg) across all tested genes; **β_D**: hurdle-model discrete component coefficient, reflecting change in the fraction of cells expressing the gene; **β_C**: continuous component coefficient, reflecting change in per-cell transcript abundance among expressing cells.

In this table, each gene is also assigned to one of four regulatory categories based on the hurdle-model coefficients β_D, β_C, and the overall log-fold-change (coef): 1) up genes show both β_D and β_C positive (and coef > 0), meaning that not only do more cells turn the gene "on," but each of those cells also produces more transcript; 2)down genes have both β_D and β_C negative (and coef < 0), reflecting that fewer cells express the gene and that those cells express it at lower levels; 3) D_driver genes exhibit a significant change in β_D (matching the sign of the coef) while β_C remains near zero or opposite in sign. The regulation occurs predominantly through recruiting more (or fewer) cells to express the gene, with little change in how much each cell expresses. When coef > 0, more cells "switch on" the gene without boosting per-cell intensity; when coef < 0, fewer cells express the gene, again without major shifts in per-cell abundance. 4) C_driver genes show the converse pattern: β_C aligns with the sign of the coef, indicating a change in transcript level per expressing cell, while β_D stays near zero or opposite in sign. This reflects modulation of expression intensity within cells, rather than changes in the number of expressing cells.

**Supplementary Table S4. MitoCarta3.0–annotated differentially expressed genes in pooled samples S1-S4 between high- and low-heteroplasmy (> 90 % vs. 0–10 %, LRT $p < 0.05$).**

| primerid | Pr(>Chisq) | coef | ci.hi | ci.lo | fdr | fold_change[1] | log2FC[2] | β_D | β_C | category |
|---|---|---|---|---|---|---|---|---|---|---|
| *ABCD3* | 0.0036 | -0.0621 | 0.0016 | -0.1258 | 0.8330 | 0.9398 | -0.0895 | -0.0441 | -0.0825 | down |
| *ACOT13* | 0.0428 | 0.0830 | 0.1569 | 0.0092 | 0.9426 | 1.0866 | 0.1198 | 0.2721 | 0.0106 | up |
| *AKR7A2* | 0.0036 | 0.0935 | 0.1565 | 0.0305 | 0.8330 | 1.0980 | 0.1349 | -0.0851 | 0.1005 | C_driver |
| *CHCHD3* | 0.0247 | -0.0722 | -0.0133 | -0.1312 | 0.9088 | 0.9303 | -0.1042 | -0.4286 | -0.0387 | down |
| *COX7A1* | 0.0049 | 0.1265 | 0.2248 | 0.0283 | 0.8645 | 1.1349 | 0.1825 | 0.3064 | -0.0033 | D_driver |
| *CYCS* | 0.0038 | -0.1125 | -0.0332 | -0.1917 | 0.8482 | 0.8936 | -0.1622 | -0.5534 | -0.0430 | down |
| *DNAJC15* | 0.0355 | 0.0038 | 0.0613 | -0.0537 | 0.9269 | 1.0038 | 0.0055 | -0.6287 | 0.0258 | C_driver |
| *EXOG* | 0.0363 | -0.0788 | -0.0161 | -0.1414 | 0.9269 | 0.9243 | -0.1136 | -0.2288 | -0.0077 | down |
| *FAM210B* | 0.0147 | -0.1055 | -0.0321 | -0.1789 | 0.9088 | 0.8999 | -0.1522 | -0.3654 | -0.0737 | down |
| *FARS2* | 0.0124 | -0.0411 | 0.0314 | -0.1135 | 0.9088 | 0.9598 | -0.0592 | 0.1140 | -0.0821 | C_driver |
| *GPX1* | 0.0095 | -0.0574 | 0.0162 | -0.1310 | 0.9088 | 0.9442 | -0.0829 | -0.4901 | -0.0262 | down |
| *HSD17B4* | 0.0362 | 0.0034 | 0.0719 | -0.0651 | 0.9269 | 1.0034 | 0.0049 | -0.1722 | 0.0550 | C_driver |
| *MGST1* | 0.0436 | 0.0241 | 0.1490 | -0.1008 | 0.9426 | 1.0244 | 0.0348 | -0.2020 | 0.0907 | C_driver |
| *MICOS10* | 0.0034 | -0.0746 | -0.0159 | -0.1333 | 0.8330 | 0.9281 | -0.1076 | -0.6297 | -0.0410 | down |
| *MRPL55* | 0.0120 | 0.0866 | 0.1532 | 0.0199 | 0.9088 | 1.0904 | 0.1249 | 0.4041 | 0.0293 | up |
| *MRPL57* | 0.0212 | 0.0719 | 0.1276 | 0.0161 | 0.9088 | 1.0745 | 0.1037 | -0.1048 | 0.0757 | C_driver |
| *MSRA* | 0.0145 | -0.0787 | -0.0075 | -0.1499 | 0.9088 | 0.9243 | -0.1135 | -0.2950 | 0.0075 | D_driver |
| *MT-ATP8* | 0.0140 | -0.0917 | -0.0270 | -0.1564 | 0.9088 | 0.9124 | -0.1323 | 0.1335 | -0.0936 | C_driver |
| *MT-ND5* | 0.0287 | -0.0805 | -0.0213 | -0.1396 | 0.9088 | 0.9227 | -0.1161 | 0.0000 | -0.0805 | down |
| *MTG2* | 0.0397 | 0.0420 | 0.1022 | -0.0182 | 0.9413 | 1.0429 | 0.0606 | 0.0425 | 0.0599 | up |
| *MTX3* | 0.0499 | -0.0481 | 0.0124 | -0.1086 | 0.9426 | 0.9531 | -0.0694 | -0.0609 | -0.0685 | down |
| *MYO19* | 0.0422 | -0.0488 | 0.0106 | -0.1082 | 0.9426 | 0.9524 | -0.0704 | -0.0631 | -0.0624 | down |
| *NDUFA6* | 0.0224 | 0.0101 | 0.0742 | -0.0541 | 0.9088 | 1.0101 | 0.0145 | -0.5727 | 0.0405 | C_driver |
| *NDUFA9* | 0.0449 | 0.0902 | 0.1613 | 0.0192 | 0.9426 | 1.0944 | 0.1302 | 0.2393 | 0.0252 | up |
| *NDUFAF7* | 0.0210 | -0.0300 | 0.0178 | -0.0778 | 0.9088 | 0.9705 | -0.0433 | -0.0353 | -0.0655 | down |
| *NDUFV3* | 0.0268 | -0.0680 | 0.0060 | -0.1421 | 0.9088 | 0.9342 | -0.0982 | -0.0258 | -0.0816 | down |
| *NOA1* | 0.0181 | 0.0518 | 0.1095 | -0.0059 | 0.9088 | 1.0531 | 0.0747 | 0.0730 | 0.0655 | up |
| *OXSM* | 0.0113 | -0.0384 | -0.0033 | -0.0735 | 0.9088 | 0.9623 | -0.0554 | -0.2752 | 0.0382 | D_driver |
| *PDE12* | 0.0315 | -0.0353 | 0.0188 | -0.0895 | 0.9222 | 0.9653 | -0.0510 | -0.0357 | -0.0642 | down |
| *PDHX* | 0.0069 | 0.0251 | 0.0826 | -0.0324 | 0.9088 | 1.0254 | 0.0362 | 0.1759 | -0.0639 | D_driver |
| *PDK3* | 0.0121 | 0.0222 | 0.0691 | -0.0247 | 0.9088 | 1.0224 | 0.0320 | 0.1864 | -0.0721 | D_driver |
| *PPTC7* | 0.0260 | -0.0680 | -0.0072 | -0.1289 | 0.9088 | 0.9342 | -0.0981 | -0.2355 | 0.0157 | D_driver |
| *SCO1* | 0.0266 | 0.0913 | 0.1594 | 0.0233 | 0.9088 | 1.0956 | 0.1318 | 0.2603 | 0.0197 | up |
| *SCO2* | 0.0470 | 0.0905 | 0.1636 | 0.0173 | 0.9426 | 1.0947 | 0.1305 | 0.2379 | 0.0366 | up |
| *SFXN1* | 0.0141 | -0.0669 | 0.0010 | -0.1349 | 0.9088 | 0.9353 | -0.0965 | -0.2474 | 0.0316 | D_driver |
| *SLC25A20* | 0.0148 | -0.0499 | -0.0058 | -0.0940 | 0.9088 | 0.9513 | -0.0720 | -0.1496 | -0.0553 | down |
| *SLC25A22* | 0.0407 | 0.0051 | 0.0320 | -0.0218 | 0.9426 | 1.0051 | 0.0074 | 0.1150 | -0.0674 | D_driver |
| *TARS2* | 0.0096 | -0.0437 | -0.0043 | -0.0832 | 0.9088 | 0.9572 | -0.0631 | -0.2578 | 0.0334 | D_driver |
| *TIMM17B* | 0.0162 | -0.0884 | -0.0134 | -0.1635 | 0.9088 | 0.9154 | -0.1276 | -0.3033 | 0.0047 | D_driver |
| *TRNT1* | 0.0059 | -0.0467 | 0.0186 | -0.1120 | 0.8856 | 0.9544 | -0.0673 | 0.0250 | -0.0812 | C_driver |
| *TXNRD2* | 0.0062 | -0.0667 | -0.0043 | -0.1290 | 0.9088 | 0.9355 | -0.0962 | -0.0698 | -0.0796 | down |
| *UQCC1* | 0.0015 | -0.0699 | -0.0085 | -0.1314 | 0.7014 | 0.9325 | -0.1009 | -0.0567 | -0.0975 | down |

[1] fold-change = (exp(coef)); [2]$\log_2$FC: base-2 log-fold-change (coef / log(2)).

### Supplementary Figure S1

**A**

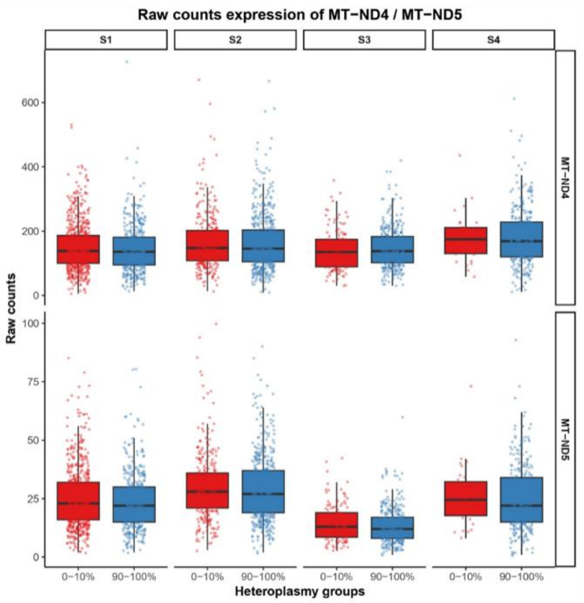


**B**

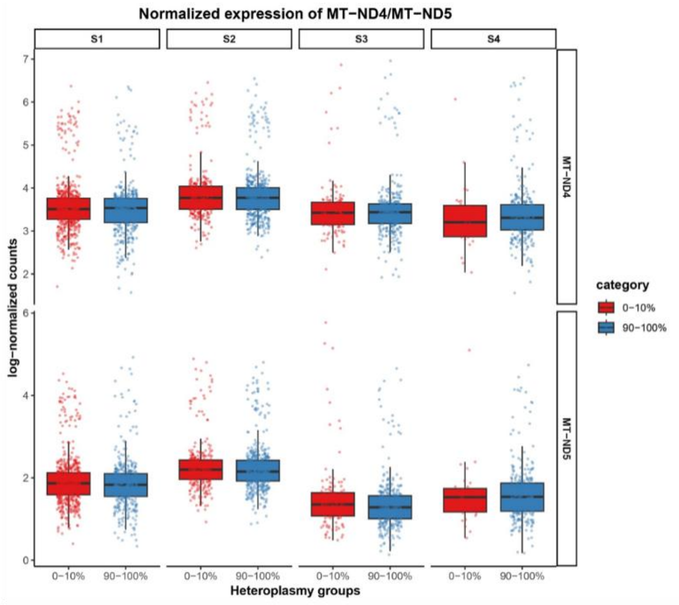


**Figure S1: Raw and log-normalized expression of mitochondrial genes MT-ND4 and MT-ND5 stratified by heteroplasmy category (0–10 % vs. 90–100 %) across four patient fibroblast samples (S1–S4).**

A) boxplots of raw UMI counts per cell, overlaid with individual data points; B) boxplots of log-normalized counts, overlaid with individual data points.

### Supplementary Figure S2

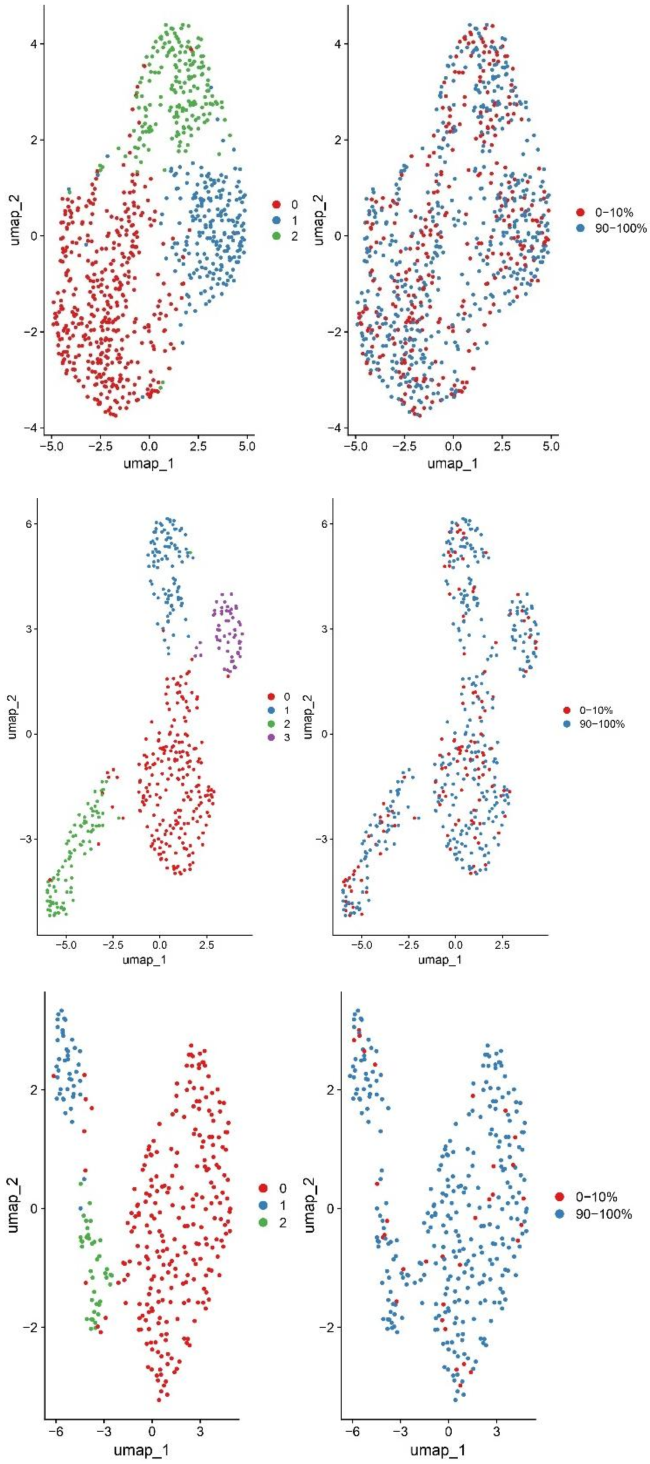


**Figure S2: Per-sample UMAP embeddings showing the distribution of low- and high-heteroplasmy cells.**

Each rows shows one fibroblast line (S2-S3-S4); for each sample we display two panels side-by-side. Left panel: UMAP embedding (resolution 0.3; graph-based clustering) with cells colored by the cluster label to visualize the local transcriptomic structure. Right panel: the same embedding colored by heteroplasmy class - low (0-10%) in blue and high (90-100%) in red - with cells that fall outside these strict thresholds shown in light gray. Only the two extreme heteroplasmy groups were contrasted to focus on putative "virtual homoplasmic" states. Across all samples the two heteroplasmy classes are extensively intermixed within clusters, mirroring the pattern shown for S1 in the main text. Note that the clarity of cluster boundaries varies between samples, but in no case do high-heteroplasmy cells form a separate global transcriptional cluster. These panels therefore support the conclusion that heteroplasmy status alone is not the primary driver of global transcriptomic variation in our fibroblast lines.
